# Comparison of the Safety and Immunogenicity of a Novel Matrix-M-adjuvanted Nanoparticle Influenza Vaccine with a Quadrivalent Seasonal Influenza Vaccine in Older Adults: A Randomized Controlled Trial

**DOI:** 10.1101/2020.08.07.20170514

**Authors:** Vivek Shinde, Iksung Cho, Joyce S. Plested, Sapeckshita Agrawal, Jamie Fiske, Rongman Cai, Haixia Zhou, Xuan Pham, Mingzhu Zhu, Shane Cloney-Clark, Nan Wang, Bin Zhou, Maggie Lewis, Patty Price-Abbott, Nita Patel, Michael J Massare, Gale Smith, Cheryl Keech, Louis Fries, Gregory M Glenn

## Abstract

**Background:** Improved seasonal influenza vaccines for older adults are urgently needed, which can induce broadly cross-reactive antibodies and enhanced T-cell responses, particularly against A(H3N2) viruses, while avoiding egg-adaptive antigenic changes.

**Methods:** We randomized 2654 clinically-stable, community-dwelling adults ≥65 years of age 1:1 to receive a single intramuscular dose of either Matrix-M-adjuvanted quadrivalent nanoparticle influenza vaccine (qNIV) or a licensed inactivated influenza vaccine (IIV4) in this randomized, observer-blinded, active-comparator controlled trial conducted during the 2019-2020 influenza season. The primary objectives were to demonstrate the non-inferior immunogenicity of qNIV relative to IIV4 against 4 vaccine-homologous strains, based on Day 28 hemagglutination-inhibiting (HAI) antibody responses, described as geometric mean titers and seroconversion rate difference between treatment groups, and to describe the safety of qNIV. Cell-mediated immune (CMI) responses were measured by intracellular cytokine analysis.

**Findings:** qNIV demonstrated immunologic non-inferiority to IIV4 against 4 vaccine-homologous strains as assessed by egg-based HAI antibody responses. Corresponding wild-type HAI antibody responses by qNIV were significantly higher than IIV4 against all 4 vaccine-homologous strains (22-66% increased) and against 6 heterologous A(H3N2) strains (34-46% increased), representing multiple genetically and/or antigenically distinct clades/subclades (all p-values <0.001). qNIV induced 3.·1- to 3·9- and 4·0- to 4·9-fold increases in various polyfunctional phenotypes of antigen-specific effector CD4+ T-cells against A(H3N2) and B/Victoria strains at Day 7 post-vaccination, respectively, while corresponding fold-rises induced by IIV4 at Day 7 were 1·3-1·4 and 1·7-2·0; representing a 126-189% improvement in CMI responses for qNIV (all p-values <0·001). Local reactogenicity, primarily mild to moderate and transient pain, was higher in the qNIV group.

**Interpretation:** qNIV was well tolerated and produced a qualitatively and quantitatively enhanced humoral and cellular immune response in older adults. These enhancements may be critical to improving the effectiveness of currently licensed influenza vaccines.

**Funding:** Novavax.

## INTRODUCTION

The substantial health and economic burden of influenza in older adults has remained largely unabated despite vaccination coverage rates in excess of 60% over the past decade, due in part to the suboptimal vaccine effectiveness (VE) of existing influenza vaccines.^1-4^ During two recent US influenza seasons, VE among older adults was reported to range between 10% and 13% for the A(H3N2) component of the vaccine—a problematic observation, because historically, A(H3N2) viruses have circulated more frequently, accounted for the majority of influenza morbidity and mortality, evolved genetically and antigenically the most rapidly, necessitating frequent vaccine strain updates, and, taken together, represented the “weak link” in influenza vaccine performance.^5-10^

Several challenges have converged over the past decade to degrade the effectiveness of traditional inactivated influenza vaccines (IIVs) in older adults.^7,8,11^ A long-standing challenge has been the induction of narrow, strain-specific antibody responses, resulting in increasing vulnerability to antigenic drift arising from an expanding diversity of circulating viruses.^11^ A second, recently recognized challenge has been the introduction of deleterious egg-adaptive antigenic changes in vaccine virus hemagglutinins (HAs), due to the near universal reliance on traditional egg-based vaccine production methods.^12-16^ A third challenge has been the limited induction of T-cell immunity by existing approaches, particularly among older adults.^17-19^

These limitations, coupled with specific characteristics of the older adult population—age-related immunosenescence, multi-morbidity, and concomitant increases in physiological frailty— have prompted the development of “enhanced” influenza vaccines, which have included a high-dose IIV (IIV-HD), a MF-59-adjuvanted IIV (aIIV), and a recombinant HA influenza vaccine (RIV).^17,20-25^ Notwithstanding, critical gaps in vaccine performance remain. IIV-HD contains a four-fold higher content of influenza antigens, while aIIV contains an oil-in-water emulsion adjuvant. Both have demonstrated improvements in hemagglutination-inhibiting (HAI) antibody responses, and in VE, compared to traditional IIVs.^23,24,26-31^ However, with both IIV-HD and aIIV, the problems of suboptimal induction of T-cell responses and introduction of egg-adaptive antigenic changes remain unaddressed; further, with IIV-HD, the improvements in efficacy have lacked breadth of cross-protection against drift variants.^18,19,24,27,32-35^ Finally, RIV, containing a three-fold higher content of influenza antigens, avoids the problems of egg-adaptive antigenic changes, and has shown improved relative VE compared to a traditional IIV during a season characterized by circulation of antigenically drift A(H3N2), but, to date, has not demonstrated substantial induction of T-cell responses.^18,25,36^

To address multiple gaps limiting the performance of existing enhanced influenza vaccines, we developed a novel recombinant, *Spodoptera frugiperda* (Sf9) insect cell/baculovirus system derived, quadrivalent HA nanoparticle influenza vaccine (qNIV), formulated with a saponin-based adjuvant, Matrix-M.^37,38^ Through previous phase 1 and 2 trials, we demonstrated that qNIV induced broadly cross-reactive HAI antibodies as compared to IIV-HD, and increased antigen-specific polyfunctional CD4+ T-cell responses as compared to IIV-HD *and* riv.^37,39,40^ To advance the candidate qNIV towards licensure via the US Food and Drug Administration’s accelerated approval pathway, we conducted a pivotal phase 3 trial to test the hypothesis that qNIV would be immunologically non-inferior to a licensed, standard-dose, quadrivalent inactivated influenza vaccine (IIV4) in older adults.

## METHODS

### Study Design

This was a phase 3, randomized, observer-blinded, active-controlled trial at 19 US clinical sites, conducted in advance of the 2019-2020 influenza season.

### Participants

Clinically stable adults aged ≥65 years who had not received an influenza vaccine within six months preceding the trial and had no known allergies or serious reactions to influenza vaccines were enrolled. Stable health was defined by absence of: a) changes in medical therapy within the preceding one month due to treatment failure or toxicity, b) medical events qualifying as serious adverse events (SAEs) within the preceding two months, and c) life-limiting diagnoses.

### Randomization and Masking

A randomization sequence was generated using an interactive web response system to allocate participants 1:1 to receive a single intramuscular dose of qNIV or IIV4. Treatment assignments were known only to the responsible unblinded vaccine administrators, who did not perform any trial assessments post-dosing. Participants and other site staff remained blinded for the duration of the trial. Stratification was by age (65 to <75 years; ≥75 years) and receipt of prior-year influenza vaccine (Supplement 1.1-1.3). This report summarizes the results of a pre-specified interim analysis, which was conducted upon completion of Day 28 visits for all participants. Blinded safety follow-up through one year is ongoing. The trial was approved by the Advarra Institutional Review Board (Columbia, Maryland, USA). All participants provided informed consent and were free to withdraw at any time from the trial (Supplement 1.4).

### Procedures

#### Vaccines

qNIV contained 60 μg recombinant hemagglutinin (HA) per strain for the four influenza strains recommended by the World Health Organization (WHO) for inclusion in the 2019-2020 Northern Hemisphere seasonal influenza vaccine, and was formulated with 75 μg of Matrix-M adjuvant. IIV4 contained 15 μg HA per recommended strain (Supplement 1.5).

#### Objectives

The primary objectives were to a) demonstrate the non-inferior immunogenicity of qNIV as compared to IIV4, in terms of HAI antibody responses assayed with classical egg-propagated virus reagents (hereafter “egg-based HAI”) to the four vaccine-homologous strains at Day 28 post-vaccination; and b) describe the safety of qNIV compared to IIV4. The secondary objective was to describe HAI antibody responses assayed with wild-type sequence HA virus-like particle (VLP) reagents (hereafter “wt-HAI”) against the four vaccine-homologous strains; multiple genetically and or antigenically distinct heterologous A(H3N2) strains; and a heterologous, antigenically distinct B strain (Figure S1). The pre-specified exploratory objective was to describe the quality and amplitude of cell-mediated immune (CMI) responses of qNIV, as measured by flow cytometry with intracellular cytokine staining analysis (ICCS) (Supplement 1.6).

### Outcomes

#### Safety

Safety was described in terms of solicited local and systemic adverse events (AEs) within seven days of vaccination, reported by participants in diaries, and as unsolicited AEs, including SAEs, medically attended adverse events (MAEs), and other significant new medical conditions (SNMCs) through Day 28 of the trial.

#### Immunogenicity

Blood samples for antibody response assessments were collected on Day 0 pre-vaccination and Day 28 post-vaccination. Peripheral blood samples for isolation of mononuclear cells (PBMCs) for CMI assessments were collected from a subset of 140 participants (~70 per treatment group) at two pre-designated sites on Day 0 pre-vaccination and Day 7 post-vaccination (Supplement 1.7).

### Statistical Analysis

The per protocol (PP) population was the primary population for immunogenicity, and included randomized participants who received the assigned dose of the test article according to the protocol, had HAI serology results at Day 0 and Day 28, and had no major protocol deviations (Figure 1). HAI antibody responses were summarized as geometric mean titers (GMTs); within group geometric mean fold-rises from pre-to post-vaccination at Day 28 (GMFR_post_/_pre_); the baseline-adjusted ratio of GMTs between qNIV and IIV4 at Day 28 (GMTR_qNIV/II_v4); seroconversion rates (SCRs); and SCR difference (SCR_qNIV_-SCR_IIV_4) (Tables 3, 4Supplement 1.8). The immunologic non-inferiority of qNIV to IIV4 was demonstrated if the *lower bound* of the two-sided 95% CI of the Day 28 post-vaccination GMTR_qNIV/IIV_4 was >0 67, *and* if the *lower bound* of the two-sided 95% CI of SCR difference at Day 28 (SCR_qNIV_-SCR_IIV_4) was ≥-10%, for all four homologous strains.

**Figure 1:**
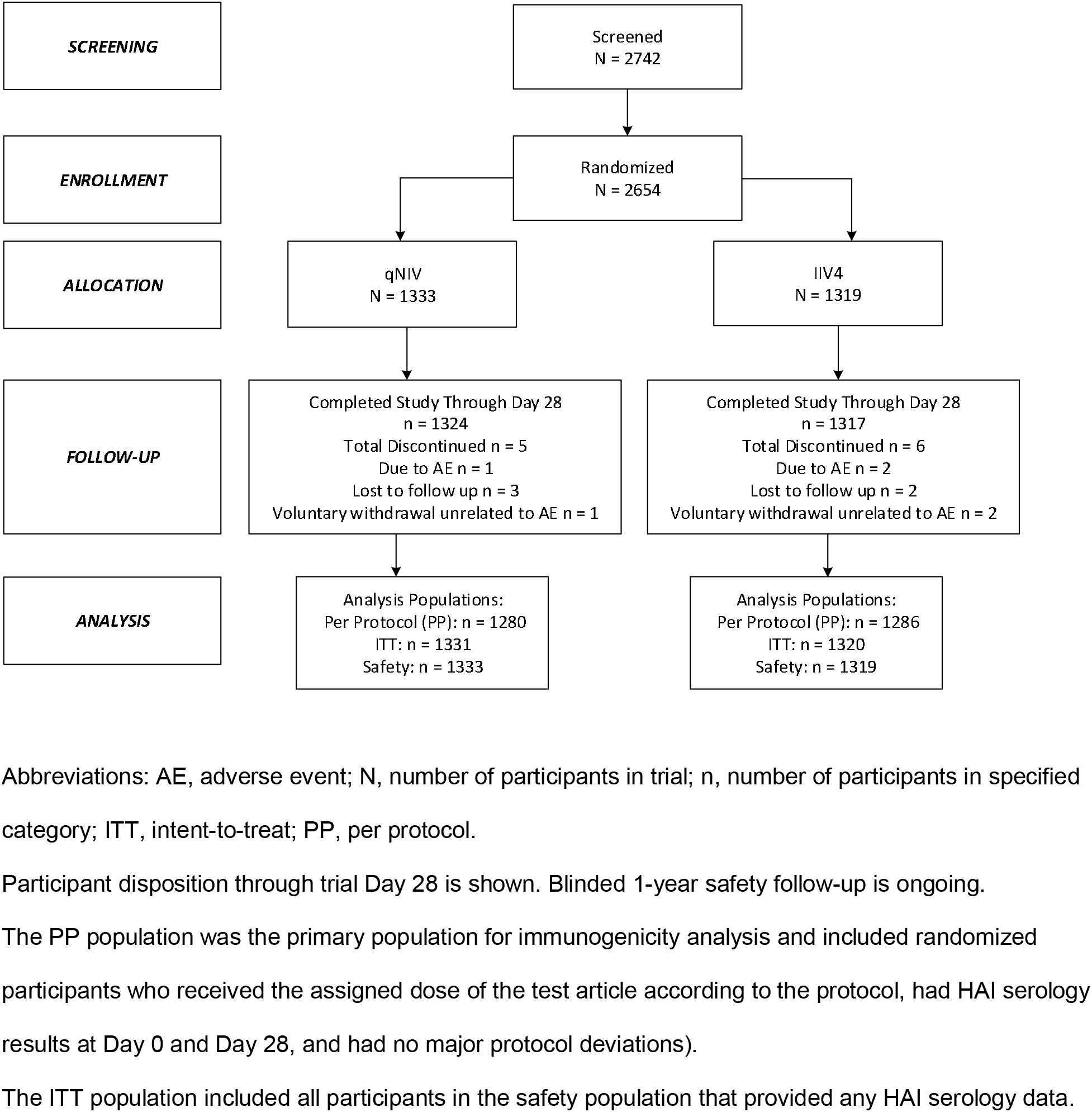
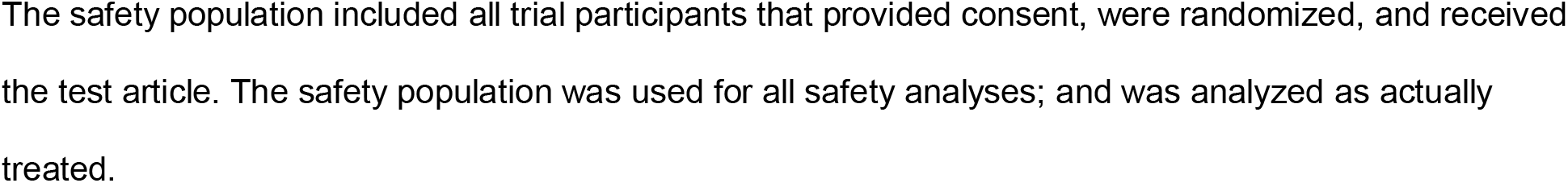
Trial Profile.

CD4+ T-cells responding to *in vitro* stimulation with homologous influenza HA antigens (A/Kansas [H3N2] or B/Maryland [Victoria]) by expressing interferon gamma (IFN-γ) either as a single cytokine marker or as polyfunctional arrays of greater than or equal to two, three, or four cytokines/activation markers consisting of combinations of the following: IFN-γ, tumor necrosis factor alpha (TNF-α), interleukin-2 (IL-2), or CD40L, which were reported as median counts and geometric mean counts (GMCs) per million cells. Within-group geometric mean fold-rises in counts from pre-to post-vaccination at Day 7 (GMFRs_post/pre_) and baseline-adjusted ratio of GMCs between qNIV and IIV4 at Day 7 (GMTR_qNIV/IIV4_) were calculated (Table S1).

The sample size of 1325 per treatment group (total 2650) was selected to achieve an overall power of 90% to demonstrate non-inferiority for all four homologous strains on both GMTR and SCR difference success criteria at a significance level of 0·025, while assuming 10% attrition (Supplement 1.8).

## RESULTS

### Participants

A total of 2654 participants were enrolled and randomized from 14-25 October, 2019, of whom 2652 received treatment on Day 0 (1333 in qNIV group and 1319 in IIV4 group) and constituted the safety population (Figure 1). The immunogenicity PP population consisted of 2566 participants. Similar percentages of participants from both treatment groups (99·3% qNIV and 99·8% IIV4) completed follow-up through Day 28. Of the 11 participants who discontinued, only one (0·1%) qNIV and two (0·2%) IIV4 participants discontinued due to an AE (Figure 1). The median participant age ranged from 71 to 72 years. The majority of participants were females (59·4% in qNIV; 64% in IIV4) and white (91%). Approximately 84% of participants in both groups received an influenza vaccine during the prior influenza season (2018-2019) (Table 1).

**Table 1:** Demographic and key baseline characteristics of trial participants in the safety population.

|  | qNIV<br>N=1333 | IIV4<br>N=1319 |
| --- | --- | --- |
| **Age** |  |  |
| Mean (SD), years | 72.5 (5.7) | 72.5 (5.7) |
| Median, years | 72.0 | 71.0 |
| **Sex** |  |  |
| Male, n (%) | 541 (40.6) | 474 (35.9) |
| Female, n (%) | 792 (59.4) | 845 (64.1) |
| **Race/Ethnicity** |  |  |
| White, n (%) | 1208 (90.6) | 1200 (91.0) |
| Black/African American, n (%) | 105 (7.9) | 103 (7.8) |
| All other, n (%) | 20 (1.5) | 16 (1.2) |
| **Received flu vaccine in 2018-2019, n (%)** | 1117 (83.8) | 1102 (83.5) |
| **Received any flu vaccine in past 3 years, n (%)** | 1210 (90.8) | 1200 (91.0) |

Abbreviations: IIV4, quadrivalent inactivated influenza vaccine; N, number of participants in treatment group; qNIV, quadrivalent nanoparticle influenza vaccine; SD, standard deviation.
The safety population included all trial participants who provided consent, were randomized, and received the test article. The safety population was used for all safety analyses and was analyzed as actually treated.

### Safety

Approximately 49·4% and 41·8% of qNIV and IIV4 participants, respectively, experienced at least one treatment-emergent adverse event (TEAE). The differences in overall TEAEs were driven primarily by differences in solicited AEs during the seven days following vaccination and, in particular, mild to moderate and transient injection site pain.

Overall solicited AEs were reported by 41·3% of qNIV and 31·8% of IIV4 participants (Table 2). Local solicited AEs followed a similar pattern (27·0% vs 18·4%), led primarily by injection site pain (25·6% vs 16·1%) and swelling (6·3% vs. 3·1%) (Table S3). Severe local solicited events were infrequent in both groups (0·6% vs 0·2%). Proportions of participants with systemic solicited AEs were comparable (27·7% vs 22·1%). The most common systemic solicited AEs were muscle pain (12·5% vs 8·0%), headache (10·7% vs 7·9%), and fatigue (9·4% vs 7·1%) (Table S3). Severe systemic solicited AEs were infrequent in both groups (1·1% vs 0·8%).

**Table 2:**
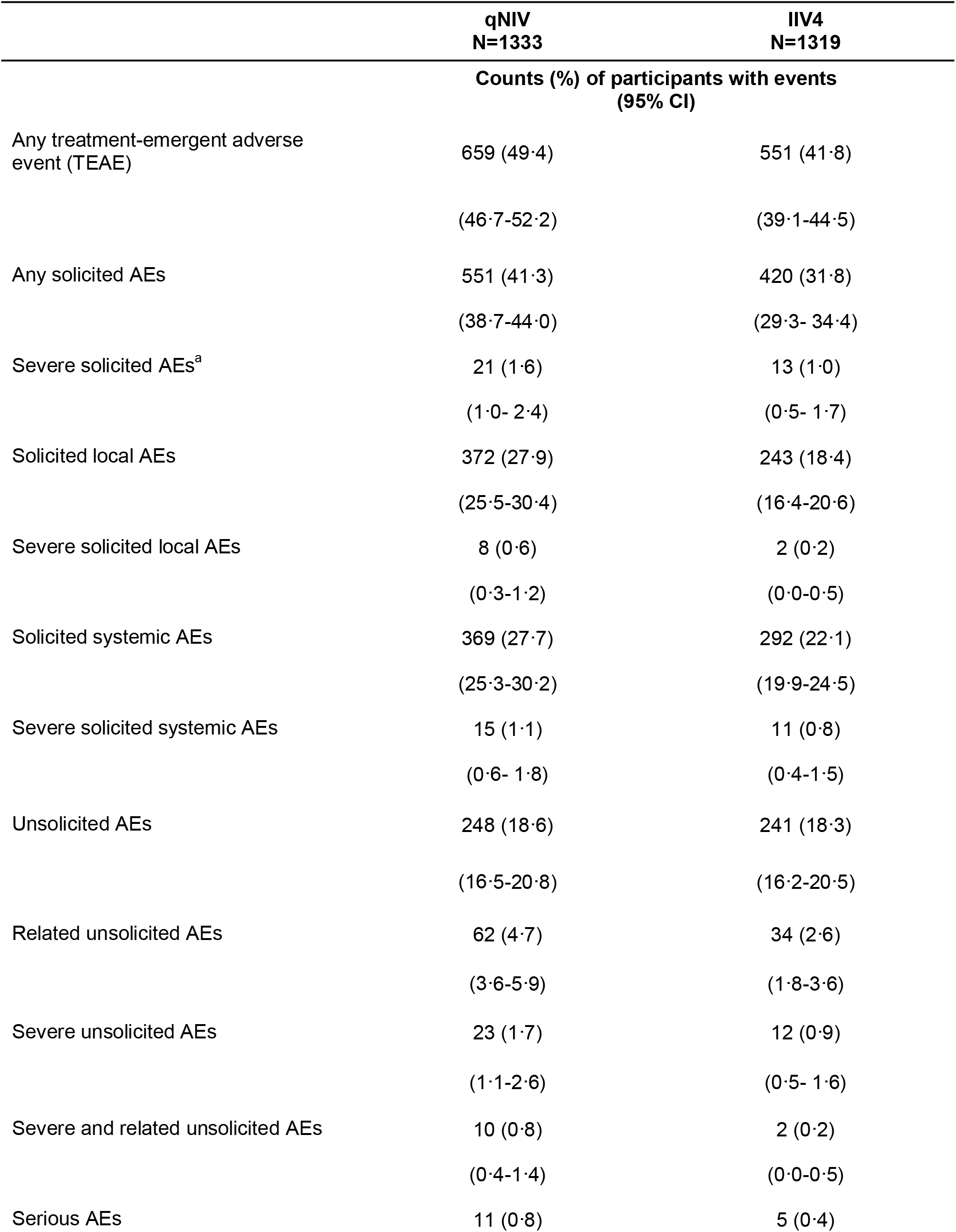
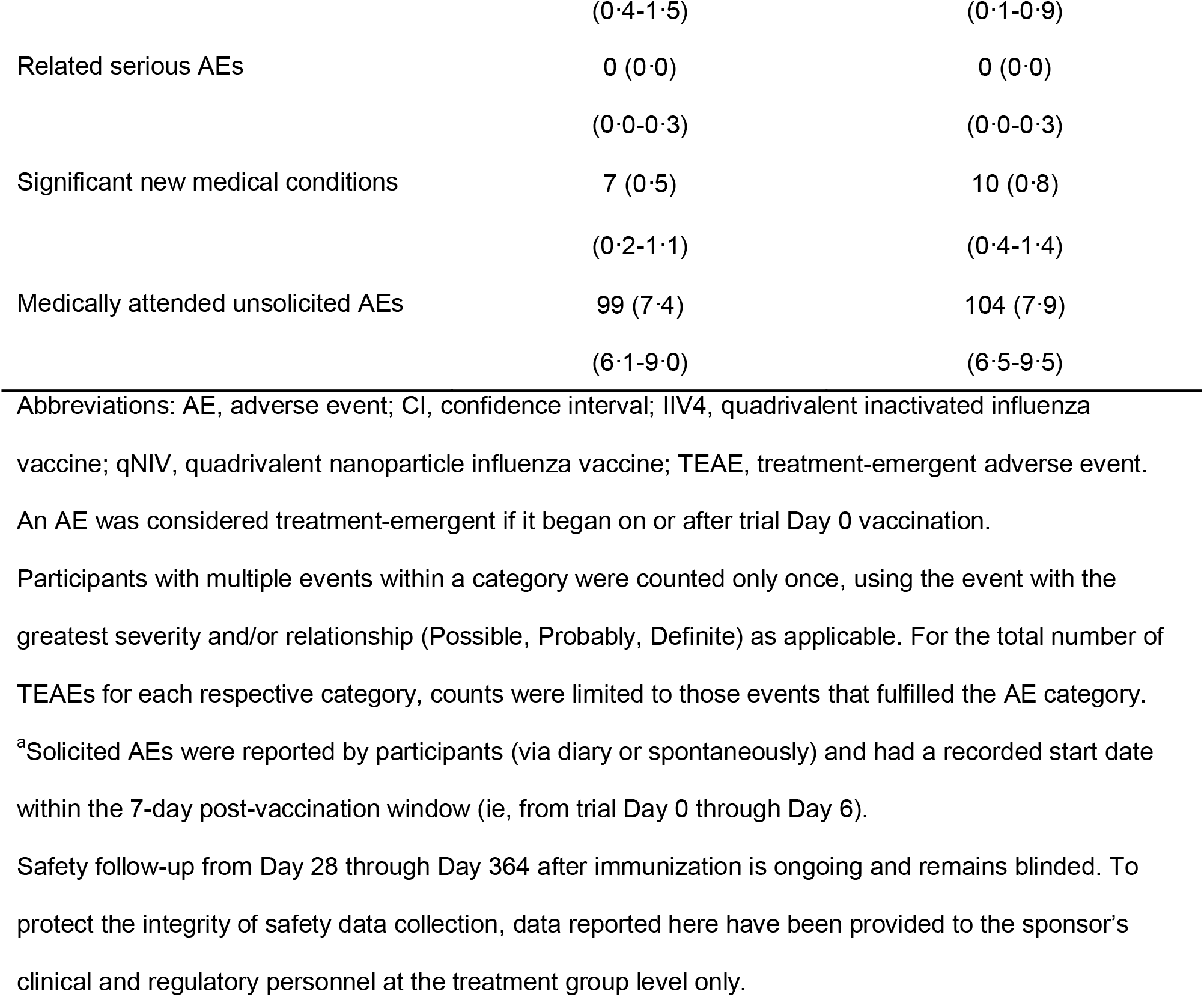
Summary of adverse events among trial participants through Day 28.

Similar proportions of participants experienced unsolicited AEs (18·6% vs 18·3%) and MAEs (7·4% vs. 7·9%) with diagnoses spanning common intercurrent illnesses for this age group, with no apparent clustering of specific AEs by treatment group. SAEs were infrequent and occurred in comparable proportions per group (0·8% vs 0·4%) (Table 2). One death occurred per treatment group; neither was considered related to treatment by trial investigators.

### Immunogenicity

#### HAI antibody responses

The primary objective of the trial was met with qNIV demonstrating immunologic non-inferiority to IIV4 against four vaccine-homologous strains based on the pre-specified GMTR and SCR difference success criteria, as assessed by egg-based HAI antibody responses, and qNIV showed statistically improved responses for three of four vaccine-homologous strains (Table 3).

**Table 3:**
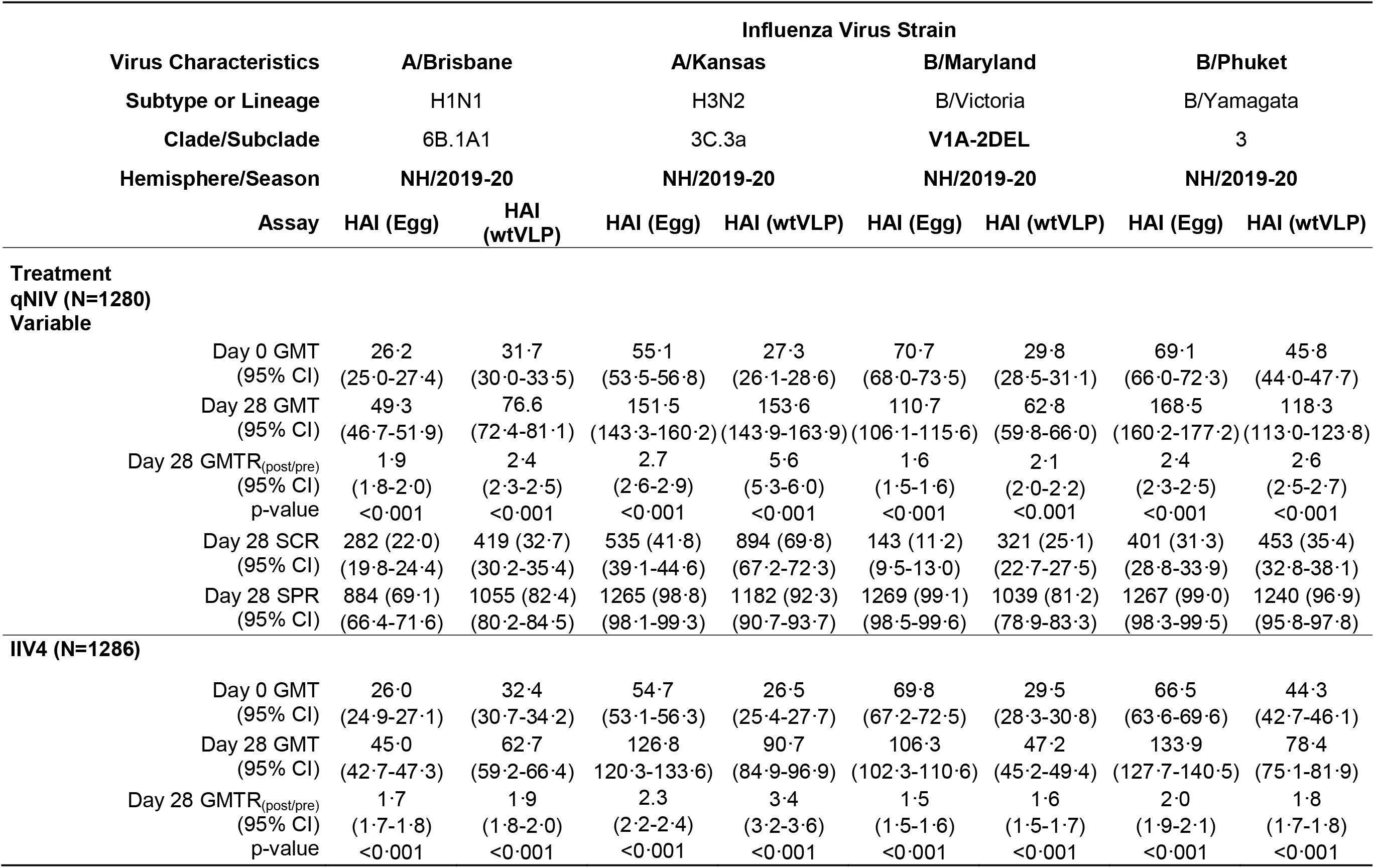
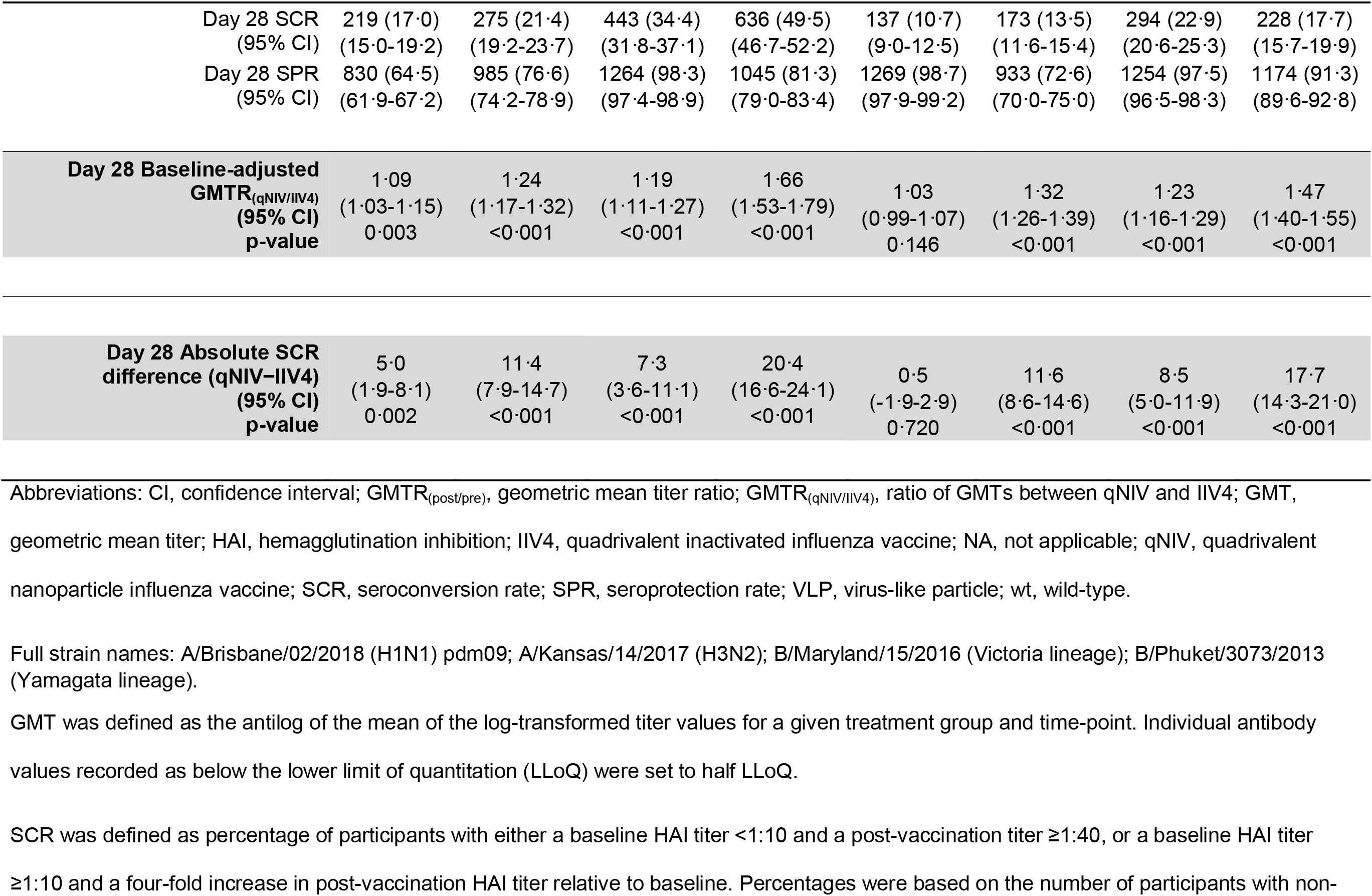
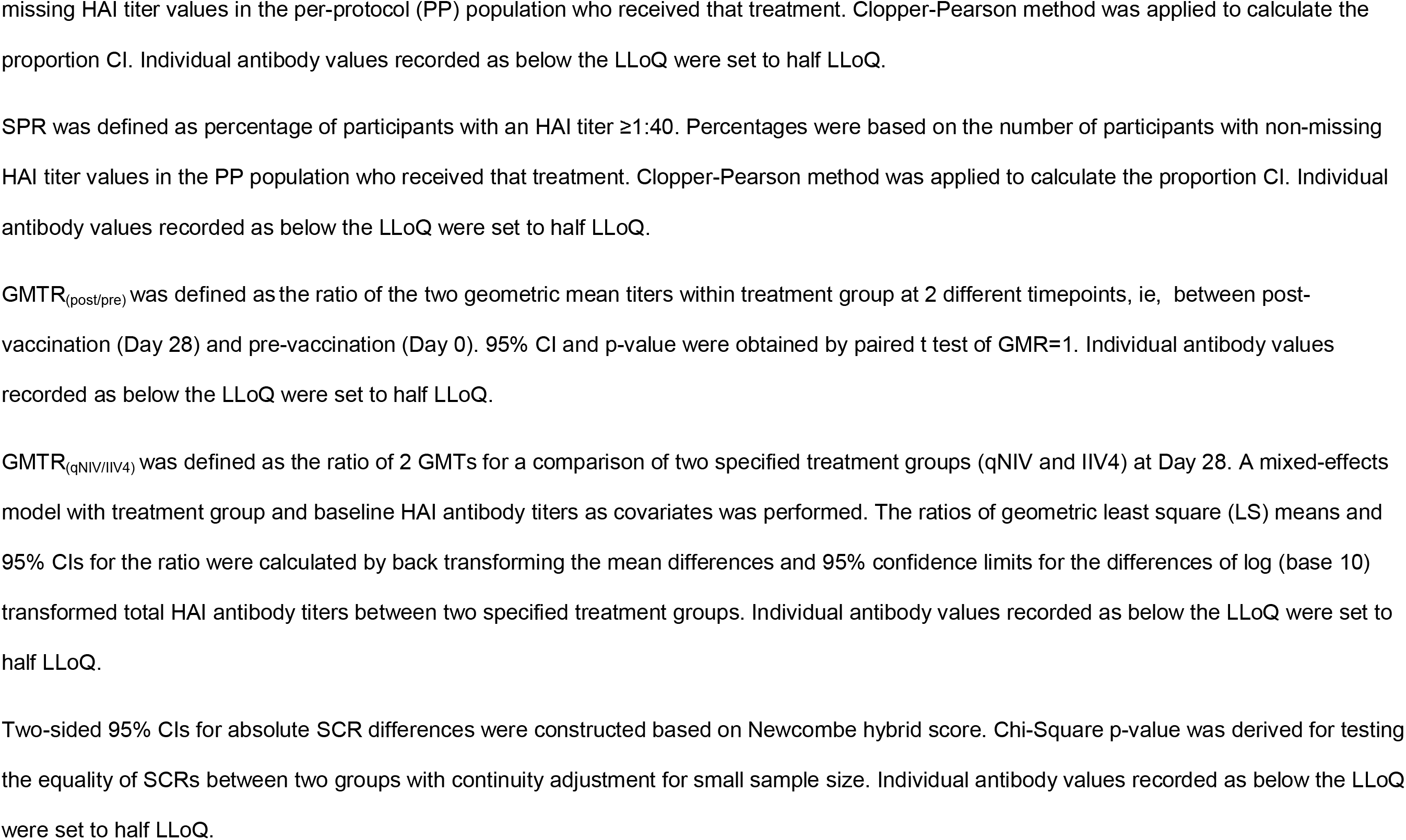
Summary of egg-virus or wild-type VLP-based Day 28 HAI GMTs, GMT ratios, SCR, and SCR difference for vaccine-homologous strains.

When HAI antibody responses were assessed in a more biologically-relevant wt-HAI assay format, which features known wild-type HA sequences and human, rather than avian, glycans as HA receptors in the agglutination reaction, the relative improvements in antibody responses were further accentuated in favor of qNIV. Specifically, Day 28 post-vaccination GMFRs for the four vaccine-homologous strains showed 2·1- to 5·6-fold increases in titers for qNIV, as compared to 1·6 -to 3·4-fold increases for IIV4. The Day 28 GMTRs showed statistically significant improvements of 24%-66% in post-vaccination wt-HAI antibody responses for qNIV as compared to IIV4 against all four vaccine-homologous strains (all p-values <0·001); corresponding Day 28 SCR differences showed increases of 11·4%-20·4% (all p-values <0·001) (Table 3).

Next, we assessed the breadth of cross-reactive antibody responses (Figure S1). GMTRs of qNIV versus IIV4 showed statistically significant improvements of 34%-46% in post-vaccination wt-HAI antibody responses against six heterologous A(H3N2) strains, and a 23% improvement against a heterologous B-Victoria lineage strain (all p-values <0·001) (Table 4).

**Table 4:**
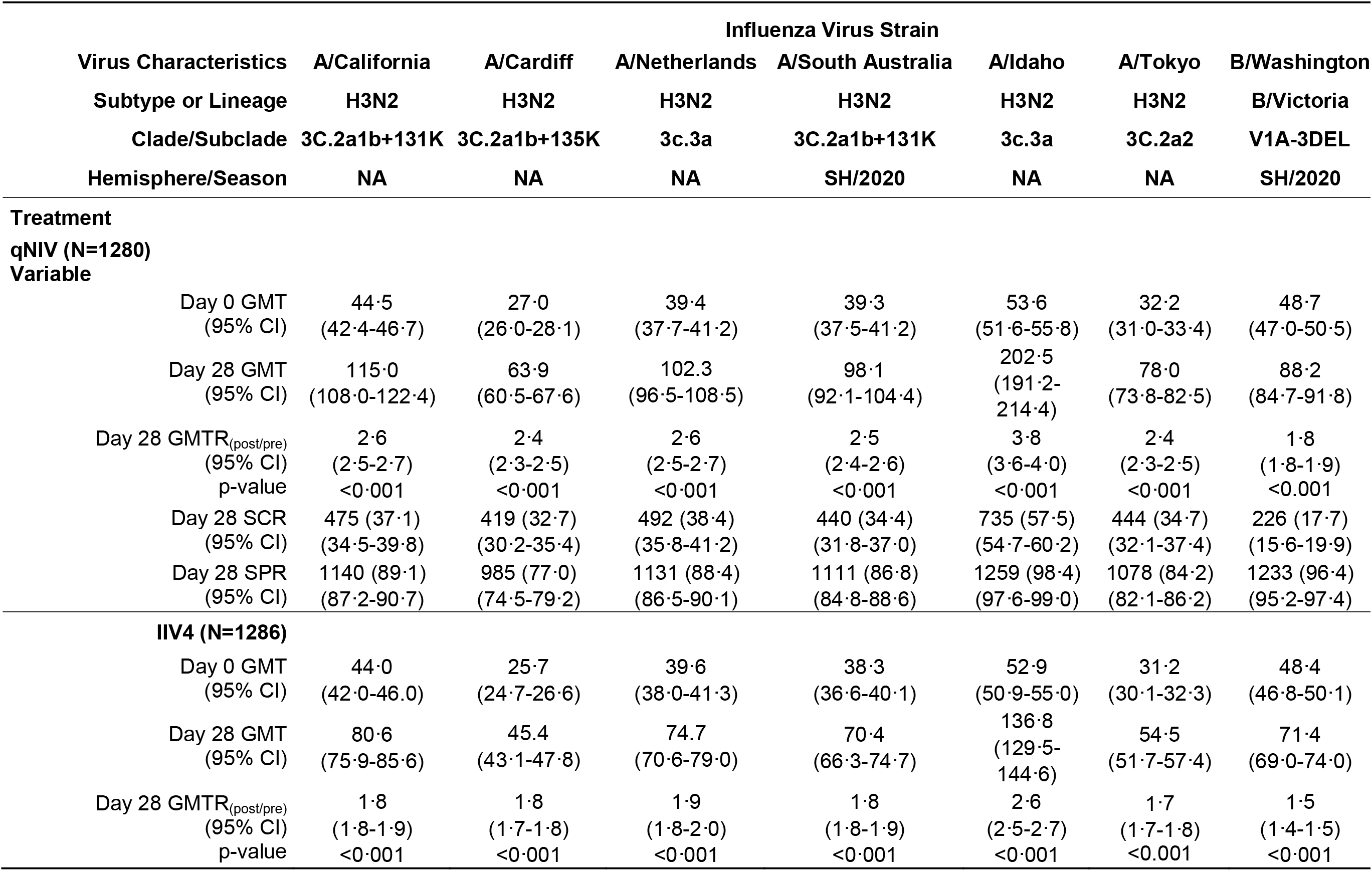
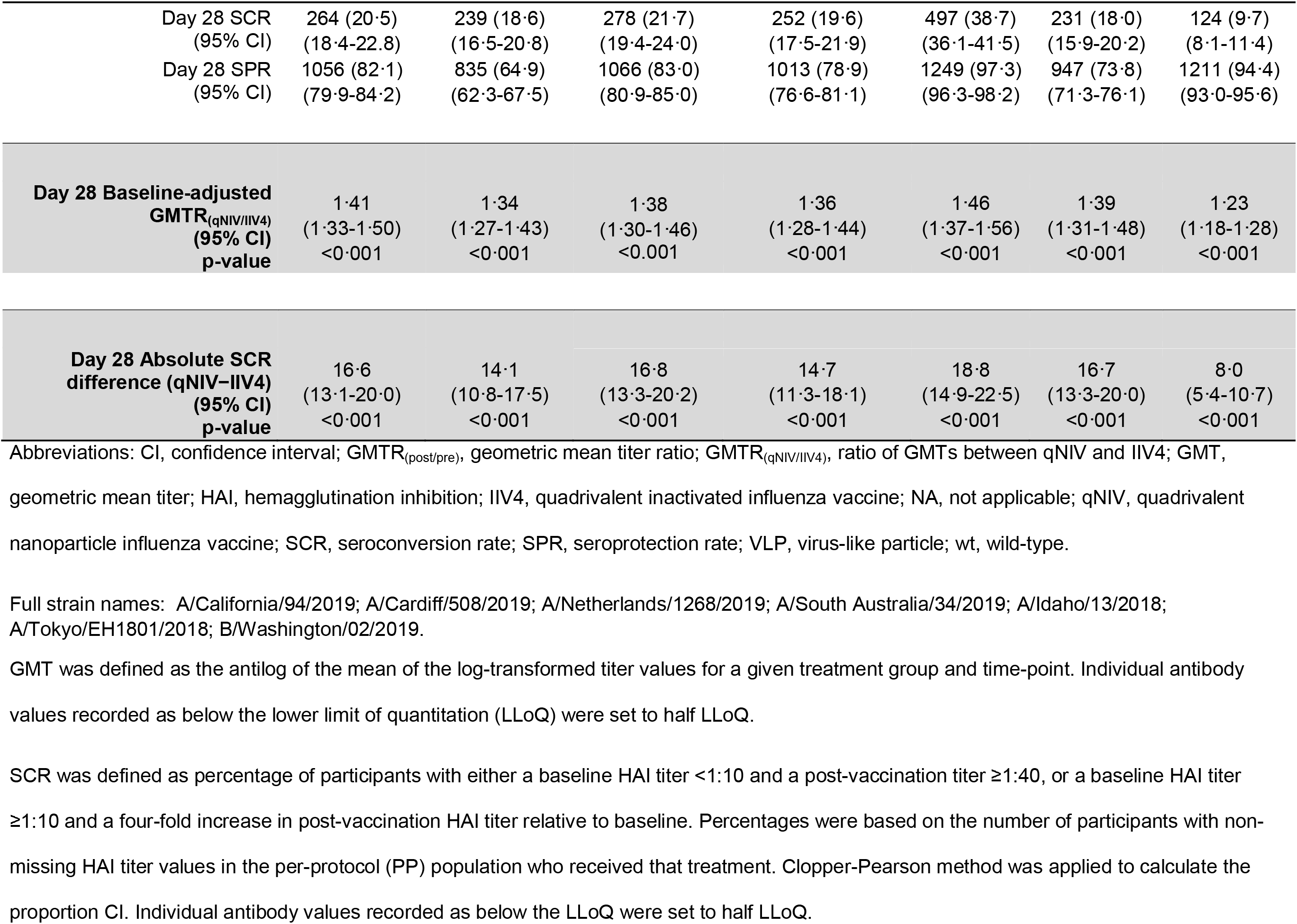
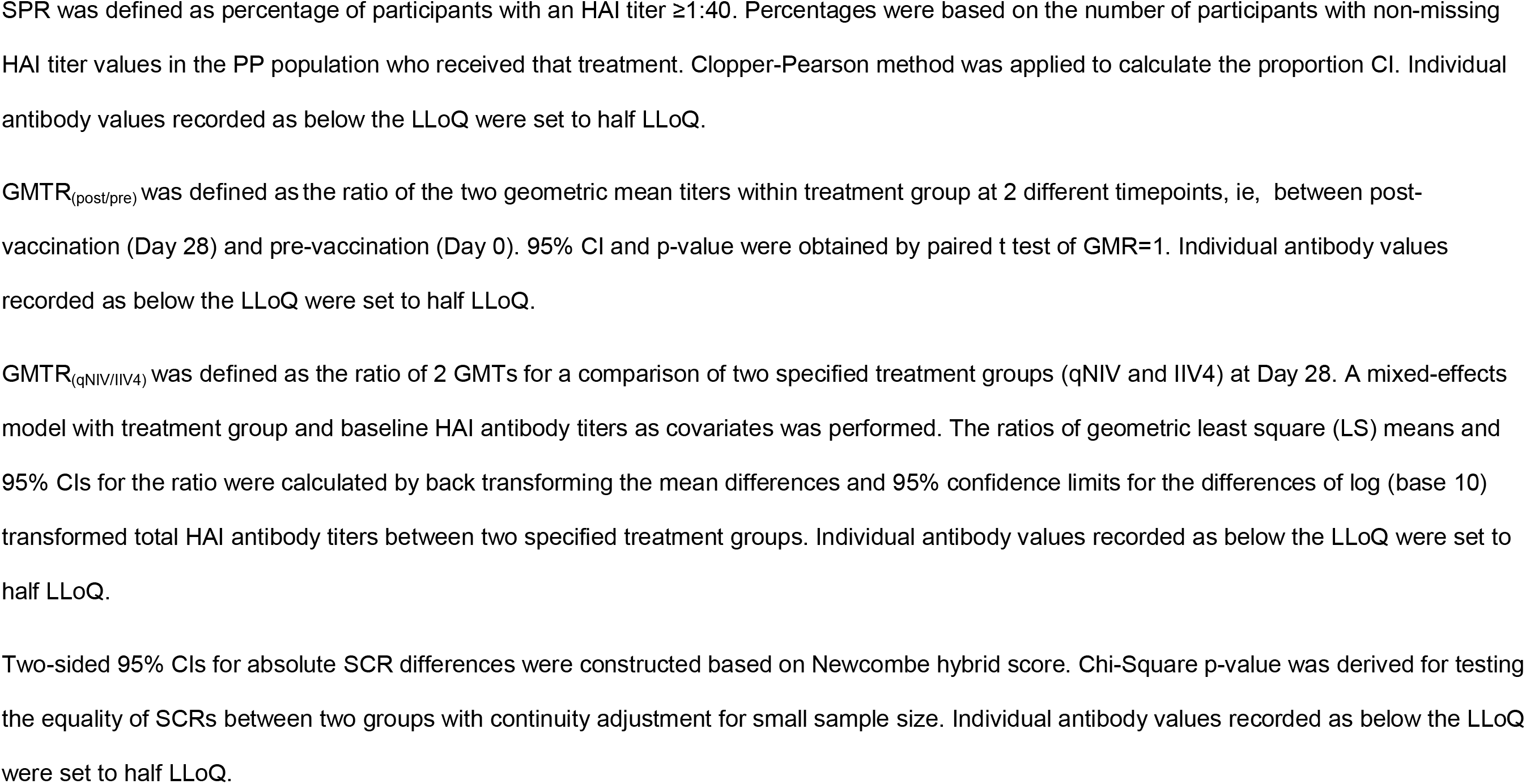
Summary of wild-type VLP-based Day 28 HAI GMTs, GMT ratios, SCR, and SCR differences for drifted A(H3N2) strains.

#### Cell-mediated immune responses

Baseline and post-vaccination CMI responses assessed across *effector* (memory) and *total* CD4+ T-cells producing single or multiple cytokines/activation markers following *in vitro* stimulation with influenza HAs are shown in Figures 2a-c, S2-S9, and Table S1. Pre-vaccination baselines were comparable in both groups. At Day 7 post-vaccination, a substantially greater induction of IFN-γ and polyfunctional responses was evident in qNIV versus IIV4 recipients, with GMFRs for qNIV stimulated with A/Kansas HA reaching 3·1-, 3·4-, 3·9-, and 4·6-fold increases for double-, triple-, quadruple-, and IFN-γ-cytokine producing *effector* CD4+ T-cells, respectively (all p-values <0·001). This was in contrast to notably lower Day 7 GMFRs observed for IIV4, ie, 1·3-, 1·3-, 1·4-, and 1·6-fold increase, respectively, for the same parameters (Figure 2c, Table S1). Corresponding GMFRs for B/Maryland followed a similar pattern (Figure 2c, Table S1). Between-group differences at Day 7, as measured by GMCR_qNIV/IIV_4, showed 141-195% relative increases in induction of double-, triple-, quadruple-, or IFN-γ-cytokine producing *effector* CD4+ T-cells following A/Kansas HA stimulation (all p-values <0·001); and 126-155% relative increases in corresponding responses after stimulation with B/Maryland HA (all p-values <0·001) (Table S1). Vaccine-induced responses measured among *total* CD4+ T-cells produced a pattern similar to effector CD4+ T-cell population for all cytokine parameters assessed (Table S1; Figures S6-S9).

**Figure 2a:**
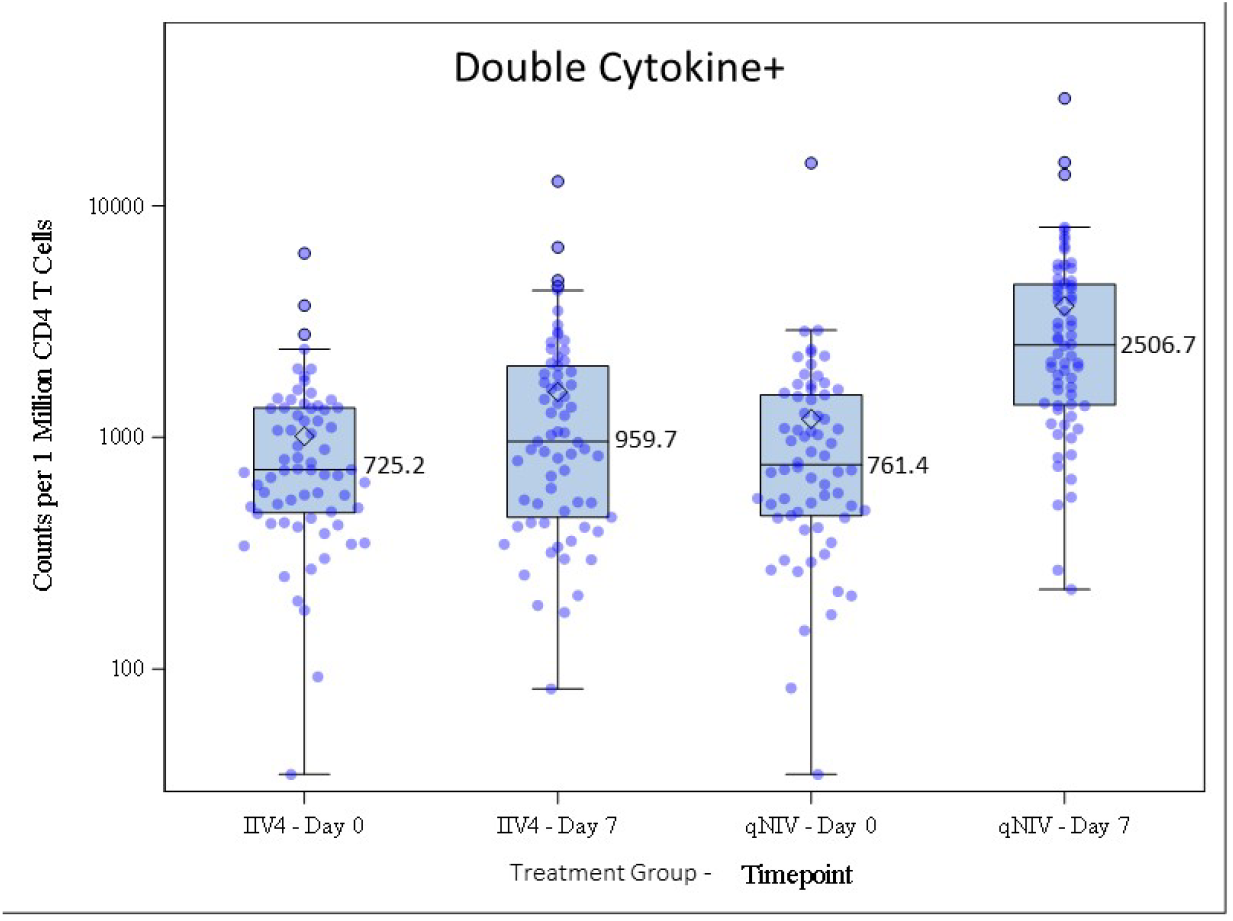
Box plot for log_10_ scale counts of at least double cytokine-expressing CD4 + effector T-cells against A/Kansas. Cell-mediated immune (CMI) responses were measured by intracellular cytokine staining (ICCS). Counts of peripheral blood CD4+T-cells expressing IL-2, IFN-γ, TNF-α and/or CD40L+ cytokines were measured following in vitro re-stimulation with A/Kansas. Responses were evaluated using peripheral blood mononuclear cells (PBMCs) obtained from a subgroup of participants on Day 0 (pre-vaccination) and Day 7. The box plots represent the interquartile range (±3 standard deviations); the solid horizontal black line represents the median and the number indicates the median count of CD4+ effector T-cells against A/Kansas, expressing at least any two of: IFN-γ, TNF-α, IL-2, or CD40L+; and the open diamond represents the mean.

**Figure 2b:**
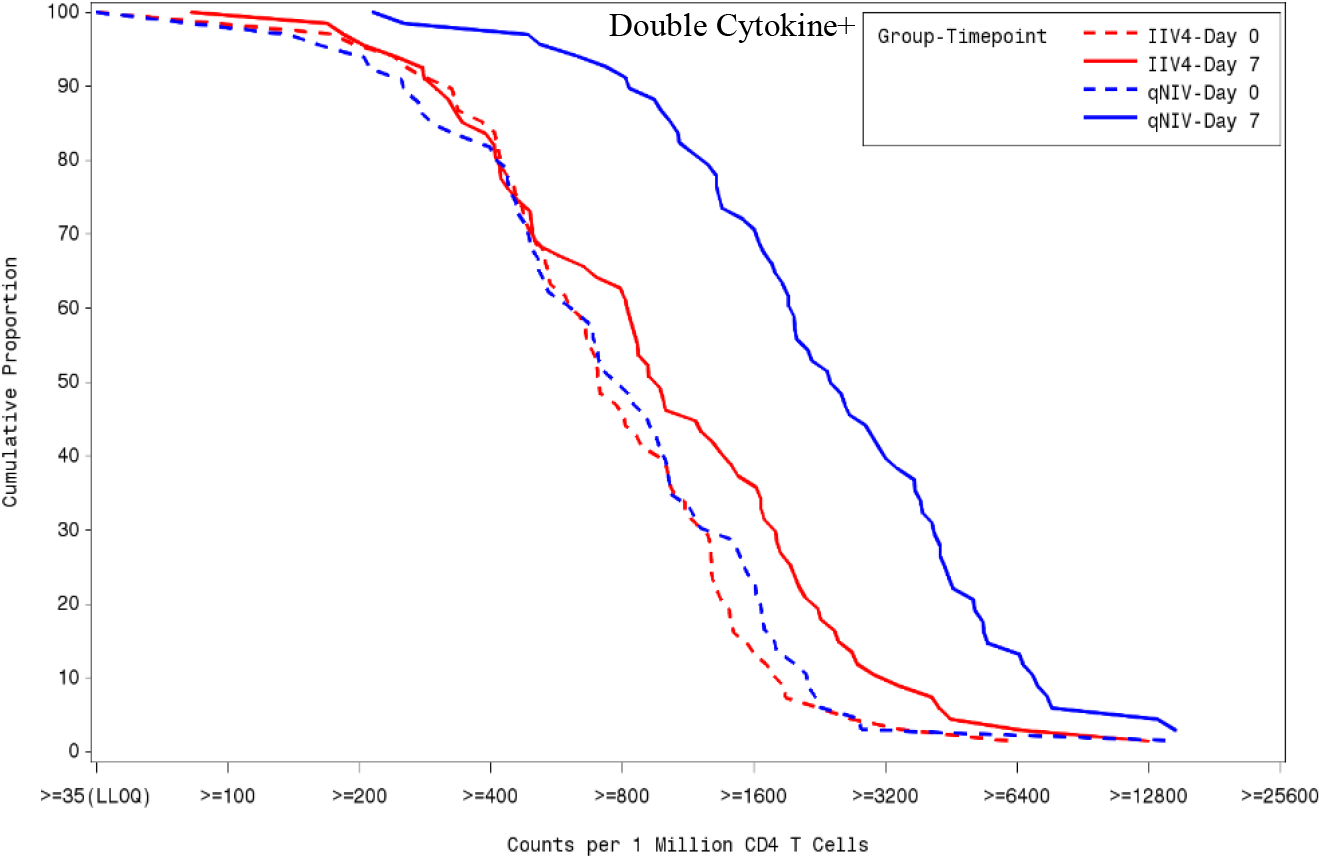
RCD plot for proportion of at least double cytokine-expressing CD4 + effector T-cells against A/Kansas.

**Figure 2c:**
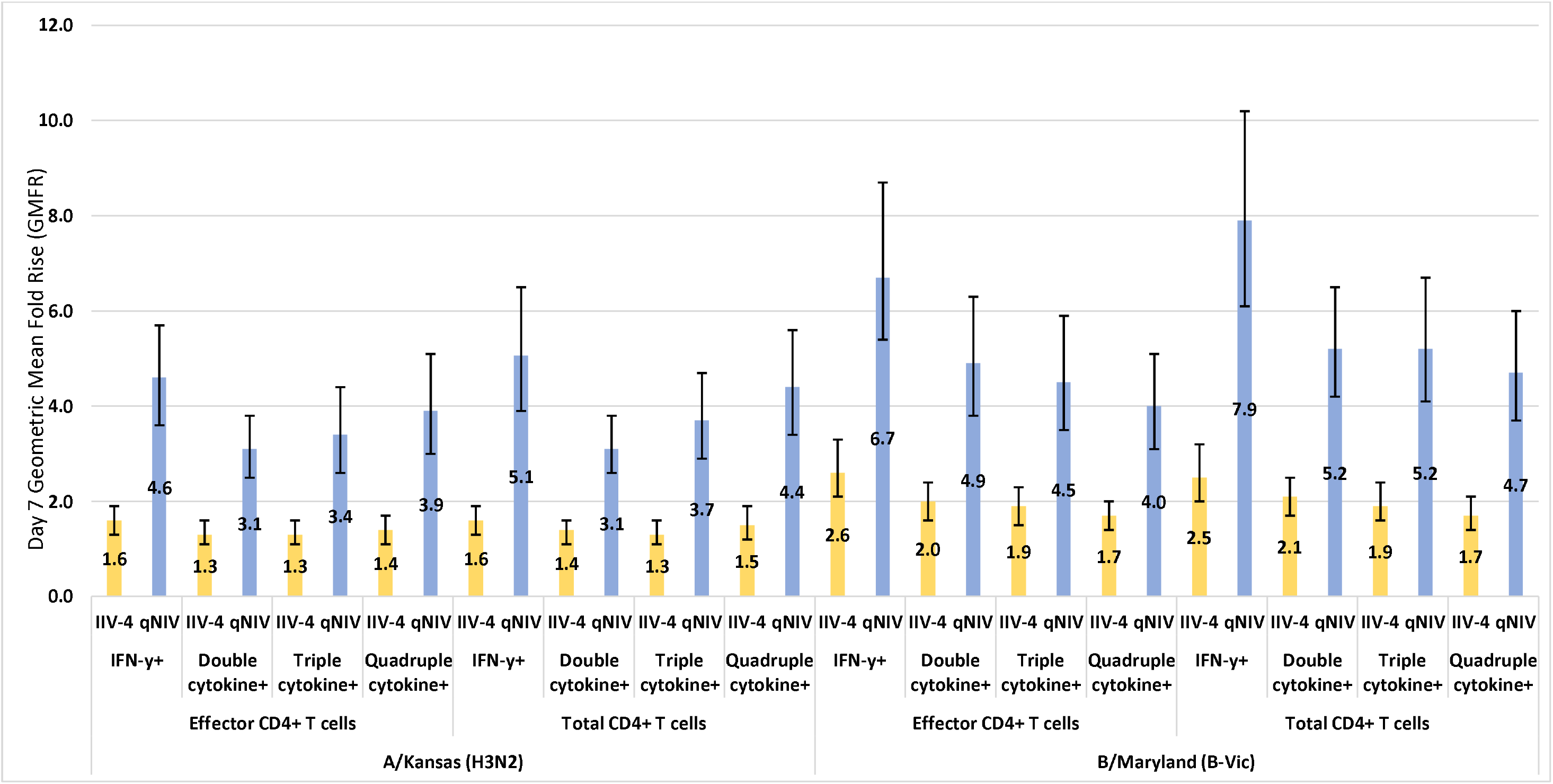
Geometric mean fold-rise at Day 7 with qNIV and IIV4 across polyfunctional phenotypes for effector and total CD4+T-cells. IFN-γ+, CD4+ effector or total T-cells expressing IFN-γ; double cytokine+, CD4+ effector or total T-610 cells expressing any two of: IFN-γ, TNF-α, IL-2, or CD40L+; triple cytokine+, CD4+ effector or total T-cells expressing any three of: IFN-γ, TNF-α, IL-2, or CD40L+; quadruple cytokine+, CD4+ effector or total T-cells expressing IFN-γ, TNF-α, IL-2, and CD40L+. Cell-mediated immune (CMI) response endpoints were performed on a subset of approximately 140 participants from several pre-designated clinical sites. For CD4+ effector T-cells, the lower limit of quantitation (LLoQ) was set as 70. If cytokine was <70, log_10_ scale of cytokines counts was recorded as log_10_ scale half LLoQ, which is 35. For CD4 + total T-cells, LLoQ was set as 40. If cytokine was <40, log_10_ scale of cytokines counts was recorded as log_10_ scale half LLoQ, which is 20. For IFN-Y, LLoQ was set as 110 for effector CD4+ T-cells, and 30 for total CD4+ T-cells. Thus, if cytokines count was <LLoQ, log_10_ scale of cytokine count was recorded as log_10_ scale half LLoQ, which is 55 and 15, respectively. GMFR_(post/pre)_ was defined as the ratio of two GMCs within treatment group at two different time-points (ie, between Day 7 and Day 0). 95% CI and p-value were obtained by paired t test of GMR=1. Note: GMC was defined as the antilog of the mean of the log transformed counts for a given treatment group and time-point.

Two overall patterns of responses were consistent across *total* and *effector* CD4+ T-cell subsets, cytokine parameters, and strains. First, as described previously, qNIV induced substantially higher post-vaccination polyfunctional CD4+ T-cell responses than IIV4 (Figures 2a-b, S2-S3, S6-S7). Second, as evident on the reverse cumulative distribution (RCD) plots, the entire distribution of pre-vaccination baseline responses were substantially and symmetrically shifted to the right following vaccination with qNIV. In contrast, in the IIV4 group, the distributions of pre-vaccination responses were more modestly and asymmetrically shifted following vaccination (Figures 2a-b, S4-S5, S8-S9). Specifically, those participants with low baseline Day 0 responses in the IIV4 group remained CMI “non-responders” to A/Kansas following vaccination; whereas, with qNIV, all participants, including those with the lowest baseline responses, achieved substantial induction of CMI responses following vaccination (Figures S4-S5,S8-S9, S11).

## DISCUSSION

In this pivotal phase 3 trial, we report four important findings. First, qNIV demonstrated non-inferiority to IIV4, based on HAI assays using conventional egg-derived reagents against the four homologous strains contained in both vaccines. Second, when HAI antibody responses were assayed with human indicator red blood cells and agglutinating reagents, which display HA proteins with wild-type sequences, a more biologically relevant assay format, qNIV induced qualitatively and quantitatively greater HAI antibody responses relative to IIV4, against both vaccine homologous strains, achieving 24-66% improvements in GMTs at Day 28, as well as against a wide array of antigenically distinct A(H3N2) strains, which showed improvements of 34-46% over IIV4 in Day 28 GMTs. Third, qNIV significantly outperformed IIV4 in the induction of various influenza HA antigen-specific polyfunctional effector (memory) and total CD4+ T-cell phenotypes, achieving increases of 126%-189% over IIV4 at Day 7 post-vaccination. Finally, qNIV was well tolerated, with a safety profile generally comparable to IIV4, except for a higher incidence of transient mild to moderate injection site pain (25·4%), which is comparable to or less than rates published for aIIV4 (24·7%) and IIV3-HD (35·3%), respectively.^41,42^

The recombinant wild-type HA antigen in qNIV is insect cell derived and forms via self-assembly of full-length HA trimers, three to seven of which further organize into a 20-40 nm sized protein-detergent nanoparticle. This format may enhance antigen recognition by increasing B- and T-cell access to conserved HA epitopes, thereby improving the quality and breadth of the antibody response.^37,38^ This was evidenced by immunization of ferrets and humans (in phase 1) with either tNIV or IIV3-HD, wherein we detected the presence of post-vaccination antibodies in tNIV treated groups that could compete with known broadly neutralizing A(H3N2)-specific monoclonal antibodies for binding to conserved HA head and stem epitopes in a manner that IIV3-HD did not, suggesting that tNIV could induce a broadly cross-reactive antibody response, while IIV-HD, by contrast, induced an antigenically constrained strain-specific response.^38,39^

Formulation of qNIV with Matrix-M adjuvant—which has been shown to enhance antigen presentation, expand the recognized antibody epitope repertoire, enhance neutralizing and cross-neutralizing antibody responses, and improve induction of potent CD4+ and CD8+T-cell responses for a variety of vaccines antigens under development—is central to improved induction of antibody and cellular responses against influenza HA antigens.^43-50^ In a previous phase 2 trial of qNIV, we showed an adjuvant effect when comparing Matrix-M-adjuvanted qNIV to unadjuvanted qNIV, demonstrating statistically significant increases in both HAI antibody (15-29% greater) and polyfunctional CD4+ T-cell (11·1-13·6 fold higher) responses.^40^ In the present trial, we again demonstrated marked induction of cell-mediated immune responses in a manner, to our knowledge, not previously reported.^18,19^ A recent randomized controlled trial in older adults from Hong Kong compared the immunogenicity of three enhanced vaccines—IIV-3 HD, aIIV, and RIV—against IIV4, and showed Day 7 post-vaccination GMFRs of IFN-γ-producing CD4+ T-cells that ranged 1·8- to 2·6-fold higher over baseline against an A(H3N2) strain for the three enhanced vaccines; and corresponding fold-rises against a B/Victoria lineage strain that ranged from 1·38 to 2·16.^18^ In contrast, in the present phase 3 trial, qNIV with Matrix-M achieved a 5·1-and 7·9-fold rise increase in Day 7 post-vaccination IFN-γ-producing CD4+ T-cell responses against A(H3N2) and B-Victoria lineage strains, respectively (Table S2). Notably, and in contrast to IIV4, qNIV with Matrix-M was able to activate influenza-specific polyfunctional cellular immune responses even among participants with the lowest levels of baseline T-cell reactivity—a crucial feature for developing protective immune responses in those with a greater degree of immunosenescence due to advanced age, physiological frailty, or multi-morbidity.^17^

Finally, the use of recombinant technology is important not only for producing a wild-type HA sequence-matched qNIV, but also for developing wild-type reagent based HAI assays to measure immunogenicity more accurately. Recent studies have elucidated the untoward effect of vaccine virus propagation in embryonated hen eggs on the emergence of deleterious antigenic site mutations on HAs of vaccine virus strains, particularly A(H3N2), which not only contribute to reduced VE due to apparent “antigenic mismatch” between circulating and egg-derived vaccine strains, but also call into question the meaningfulness of traditional HAI antibody measurements assayed with egg-propagated reagents derived in the same fashion.^8,12,13,15,16^ Our phase 2 and 3 data further underscore this problem: in the phase 3 trial, we noted a substantial improvement in the relative HAI antibody responses comparing qNIV and IIV4 when assayed with egg-based reagents (3-23% relative improvement) versus when assayed with wild-type reagents (24-66% relative improvement) (Table 3), and in phase 2 trial, we could demonstrate orthogonal support for the discrepant egg-based versus wild-type HAI observations by comparing the performance of microneutralization antibody assays employing egg-derived versus wild-type virus reagents (Figure S10).^40^

Our findings indicate that the improved humoral and cellular immune responses elicited by qNIV—a consequence of the nanoparticle antigen structure, the Matrix-M adjuvant, and recombinant technology—hold potential to address critical gaps in currently licensed influenza vaccines.

## Data Availability

The datasets generated during and/or analyzed during the current study are not publicly available as analyses are ongoing and will be used for regulatory submissions for the vaccine described; but may become available in the future from the corresponding author on reasonable request

## CONTRIBUTORS

All co-authors contributed substantially to the conception and development of the trial. VS, LF, GG, CK, and IC contributed substantially to the design and interpretation of data, and manuscript review. IC provided statistical expertise, developed the statistical analysis plan, and source tables, listings, and figures. RC and NW provided statistical programming. JP, MZ, BZ and SC developed and conducted HAI assays. BZ conducted nonclinical assays to support scientific development of trial design. HZ developed and conducted CMI assays. GS, NP, MM provided scientific expertise for the development of the study and manuscript concepts. SA drafted the manuscript and developed manuscript tables and figures. JF, ML, PP provided clinical operations, regulatory, and pharmacovigilance support. XP provided medical writing support for trial protocol and amendments.

## ACKNOWLEDGMENTS

We thank the trial participants, the investigators from the clinical trial sites, members of the sponsor’s and the contract research organization (CRO)’s team, and the Clinical Immunology laboratory for their contributions to the trial. Editorial assistance on the preparation of this manuscript was provided by Phase Five Communications, supported by Novavax, Inc.

## Clinicaltrials.gov Registration

NCT04120194

## Role of the Funder/Supporter

The Sponsor funded the trial and was responsible for the design and conduct of the study; collection, management, analysis, and interpretation of the data; preparation, review, and approval of the manuscript; and decision to submit the manuscript for publication.

## References

1. CDC. Vaccination Coverage among Adults in the United States, National Health Interview Survey, 2017. https://www.cdc.gov/vaccines/imz-managers/coverage/adultvaxview/pubs-resources/NHIS-2017.html (accessed May 02, 2020 2020).

2. Reed C, Chaves SS, Daily Kirley P, et al. Estimating Influenza Disease Burden from Population-Based Surveillance Data in the United States. PLOS ONE 2015; 10(3): e0118369.

3. Grohskopf LA, Alyanak E, Broder KR, Walter EB, Fry AM, Jernigan DB. Prevention and Control of Seasonal Influenza with Vaccines: Recommendations of the Advisory Committee on Immunization Practices — United States, 2019-20 Influenza Season. MMWR Recommendations and Reports 2019; 68(3): 1–21.

4. CDC) UCfDCaPU. Past Seasons Vaccine Effectiveness Estimates. https://www.cdc.gov/flu/vaccines-work/past-seasons-estimates.html (accessed July 26, 2020.

5. Flannery B, Kondor RJG, Chung JR, et al. Spread of Antigenically Drifted Influenza A(H3N2) Viruses and Vaccine Effectiveness in the United States During the 2018-2019 Season. J Infect Dis 2020; 221(1): 8–15.

6. Rolfes MA, Flannery B, Chung JR, et al. Effects of Influenza Vaccination in the United States During the 2017-2018 Influenza Season. Clin Infect Dis 2019; 69(11): 1845–53.

7. Belongia EA, McLean HQ. Influenza Vaccine Effectiveness: Defining the H3N2 Problem. Clin Infect Dis 2019; 69(10): 1817–23.

8. Paules CI, Sullivan SG, Subbarao K, Fauci AS. Chasing Seasonal Influenza - The Need for a Universal Influenza Vaccine. N Engl J Med 2018; 378(1): 7–9.

9. Matias G, Taylor R, Haguinet F, Schuck-Paim C, Lustig R, Shinde V. Estimates of mortality attributable to influenza and RSV in the United States during 1997-2009 by influenza type or subtype, age, cause of death, and risk status. Influenza and Other Respiratory Viruses 2014; 8(5): 507–15.

10. Matias G, Taylor R, Haguinet F, Schuck-Paim C, Lustig R, Shinde V. Estimates of hospitalization attributable to influenza and RSV in the US during 1997-2009, by age and risk status. BMC Public Health 2017; 17(1).

11. Paules CI, Fauci AS. Influenza Vaccines: Good, but We Can Do Better. The Journal of Infectious Diseases 2019; 219(Supplement_1): S1–S4.

12. Zost SJ, Parkhouse K, Gumina ME, et al. Contemporary H3N2 influenza viruses have a glycosylation site that alters binding of antibodies elicited by egg-adapted vaccine strains. Proc Natl Acad Sci U S A 2017; 114(47): 12578–83.

13. Gouma S, Zost SJ, Parkhouse K, et al. Comparison of human H3N2 antibody responses elicited by egg-based, cell-based, and recombinant protein-based influenza vaccines during the 2017-2018 season. Clin Infect Dis 2019.

14. Paules CI, Fauci AS. Influenza Vaccines: Good, but We Can Do Better. J Infect Dis 2019; 219(Suppl_1): S1–S4.

15. Wu NC, Zost SJ, Thompson AJ, et al. A structural explanation for the low effectiveness of the seasonal influenza H3N2 vaccine. PLOS Pathogens 2017; 13(10): e1006682.

16. Levine MZ, Martin ET, Petrie JG, et al. Antibodies Against Egg- and Cell-Grown Influenza A(H3N2) Viruses in Adults Hospitalized During the 2017-2018 Influenza Season. The Journal of Infectious Diseases 2019; 219(12): 1904–12.

17. McElhaney JE, Andrew MK, Haynes L, Kuchel GA, McNeil SA, Pawelec G. Influenza Vaccination: Accelerating the Process for New Vaccine Development in Older Adults. Interdiscip Top Gerontol Geriatr 2020; 43: 98–112.

18. Cowling BJ, Perera R, Valkenburg SA, et al. Comparative Immunogenicity of Several Enhanced Influenza Vaccine Options for Older Adults: A Randomized, Controlled Trial. Clin Infect Dis 2019.

19. Kumar A, McElhaney JE, Walrond L, et al. Cellular immune responses of older adults to four influenza vaccines: Results of a randomized, controlled comparison. Human Vaccines & Immunotherapeutics 2017; 13(9): 2048–57.

20. McElhaney JE, Zhou X, Talbot HK, et al. The unmet need in the elderly: how immunosenescence, CMV infection, co-morbidities and frailty are a challenge for the development of more effective influenza vaccines. Vaccine 2012; 30(12): 2060–7.

21. DiazGranados CA, Dunning AJ, Kimmel M, et al. Efficacy of high-dose versus standard-dose influenza vaccine in older adults. N Engl J Med 2014; 371(7): 635–45.

22. Izikson R, Leffell DJ, Bock SA, et al. Randomized comparison of the safety of Flublok(®) versus licensed inactivated influenza vaccine in healthy, medically stable adults ≥ 50 years of age. Vaccine 2015; 33(48): 6622–8.

23. Frey SE, Reyes MR, Reynales H, et al. Comparison of the safety and immunogenicity of an MF59(R)-adjuvanted with a non-adjuvanted seasonal influenza vaccine in elderly subjects. Vaccine 2014; 32(39): 5027–34.

24. Diazgranados CA, Dunning AJ, Kimmel M, et al. Efficacy of High-Dose versus Standard-Dose Influenza Vaccine in Older Adults. New England Journal of Medicine 2014; 371(7): 635–45.

25. Dunkle LM, Izikson R, Patriarca P, et al. Efficacy of Recombinant Influenza Vaccine in Adults 50 Years of Age or Older. N Engl J Med 2017; 376(25): 2427–36.

26. Chang LJ, Meng Y, Janosczyk H, Landolfi V, Talbot HK, Group QS. Safety and immunogenicity of high-dose quadrivalent influenza vaccine in adults ≥65_years of age: A phase 3 randomized clinical trial. Vaccine 2019; 37(39): 5825–34.

27. Tsang P, Gorse GJ, Strout CB, et al. Immunogenicity and safety of Fluzone(®) intradermal and high-dose influenza vaccines in older adults ≥65 years of age: a randomized, controlled, phase II trial. Vaccine 2014; 32(21): 2507–17.

28. Essink B, Fierro C, Rosen J, et al. Immunogenicity and safety of MF59-adjuvanted quadrivalent influenza vaccine versus standard and alternate B strain MF59-adjuvanted trivalent influenza vaccines in older adults. Vaccine 2020; 38(2): 242–50.

29. Belongia EA, Sundaram ME, McClure DL, Meece JK, Ferdinands J, Vanwormer JJ. Waning vaccine protection against influenza A (H3N2) illness in children and older adults during a single season. Vaccine 2015; 33(1): 246–51.

30. Van Buynder PG, Konrad S, Van Buynder JL, et al. The comparative effectiveness of adjuvanted and unadjuvanted trivalent inactivated influenza vaccine (TIV) in the elderly. Vaccine 2013; 31(51): 6122–8.

31. Mannino S, Villa M, Apolone G, et al. Effectiveness of Adjuvanted Influenza Vaccination in Elderly Subjects in Northern Italy. American Journal of Epidemiology 2012; 176(6): 527–33.

32. Tsai TF. Fluad(R)-MF59(R)-Adjuvanted Influenza Vaccine in Older Adults. Infect Chemother 2013; 45(2): 159–74.

33. Yoo BW, Kim CO, Izu A, Arora AK, Heijnen E. Phase 4, Post-Marketing Safety Surveillance of the MF59-Adjuvanted Influenza Vaccines FLUAD(R) and VANTAFLU(R) in South Korean Subjects Aged >/=65 Years. Infect Chemother 2018; 50(4): 301–10.

34. Frey S, Poland G, Percell S, Podda A. Comparison of the safety, tolerability, and immunogenicity of a MF59-adjuvanted influenza vaccine and a non-adjuvanted influenza vaccine in non-elderly adults. Vaccine 2003; 21(27-30): 4234–7.

35. Izurieta HS, Chillarige Y, Kelman J, et al. Relative Effectiveness of Cell-Cultured and Egg-Based Influenza Vaccines Among Elderly Persons in the United States, 2017-2018. The Journal of Infectious Diseases 2019; 220(8): 1255–64.

36. Baxter R, Patriarca PA, Ensor K, Izikson R, Goldenthal KL, Cox MM. Evaluation of the safety, reactogenicity and immunogenicity of FluBlok® trivalent recombinant baculovirus-expressed hemagglutinin influenza vaccine administered intramuscularly to healthy adults 50-64 years of age. Vaccine 2011; 29(12): 2272–8.

37. Smith G, Liu Y, Flyer D, et al. Novel hemagglutinin nanoparticle influenza vaccine with Matrix-M adjuvant induces hemagglutination inhibition, neutralizing, and protective responses in ferrets against homologous and drifted A(H3N2) subtypes. Vaccine 2017; 35(40): 5366–72.

38. Portnoff AD, Patel N, Massare MJ, et al. Influenza Hemagglutinin Nanoparticle Vaccine Elicits Broadly Neutralizing Antibodies against Structurally Distinct Domains of H3N2 HA. Vaccines 2020; 8(1): 99.

39. Shinde V, Fries L, Wu Y, et al. Improved Titers against Influenza Drift Variants with a Nanoparticle Vaccine. N Engl J Med 2018; 378(24): 2346–8.

40. Shinde V. Induction of Cross-reactive Hemagglutination Inhibiting Antibody and Polyfunctional CD4+ T-cell Responses by a Recombinant Matrix-M-Adjuvanted Hemagglutinin Nanoparticle Influenza Vaccine medRxiv 2020.

41. Sanofi Pasteur. Fluzone® High-Dose (Influenza Vaccine) [package insert]. U.S. Food and Drug Administration. https://www.fda.gov/media/119870/download (accessed June 2, 2020.

42. Seqirus. FLUAD® (Influenza Vaccine, Adjuvanted) [package insert]. U.S. Food and Drug Administration. https://www.fda.gov/media/94583/download (accessed June 2, 2020.

43. Bengtsson KL, Karlsson KH, Magnusson SE, Reimer JM, Stertman L. Matrix-M adjuvant: enhancing immune responses by ‘setting the stage’ for the antigen. Expert Rev Vaccines 2013; 12(8): 821–3.

44. Bengtsson KL, Song H, Stertman L, et al. Matrix-M adjuvant enhances antibody, cellular and protective immune responses of a Zaire Ebola/Makona virus glycoprotein (GP) nanoparticle vaccine in mice. Vaccine 2016; 34(16): 1927–35.

45. Cox RJ, Pedersen G, Madhun AS, et al. Evaluation of a virosomal H5N1 vaccine formulated with Matrix M adjuvant in a phase I clinical trial. Vaccine 2011; 29(45): 8049–59.

46. Magnusson SE, Altenburg AF, Bengtsson KL, et al. Matrix-M adjuvant enhances immunogenicity of both protein- and modified vaccinia virus Ankara-based influenza vaccines in mice. Immunol Res 2018; 66(2): 224–33.

47. Magnusson SE, Reimer JM, Karlsson KH, Lilja L, Bengtsson KL, Stertman L. Immune enhancing properties of the novel Matrix-M adjuvant leads to potentiated immune responses to an influenza vaccine in mice. Vaccine 2013; 31(13): 1725–33.

48. Reimer JM, Karlsson KH, Lovgren-Bengtsson K, Magnusson SE, Fuentes A, Stertman L. Matrix-M adjuvant induces local recruitment, activation and maturation of central immune cells in absence of antigen. PLoS One 2012; 7(7): e41451.

49. Fries L, Cho I, Krähling V, et al. Randomized, Blinded, Dose-Ranging Trial of an Ebola Virus Glycoprotein Nanoparticle Vaccine With Matrix-M Adjuvant in Healthy Adults. The Journal of Infectious Diseases 2019.

50. Fries LF, Smith GE, Glenn GM. A Recombinant Viruslike Particle Influenza A (H7N9) Vaccine. New England Journal of Medicine 2013; 369(26): 2564–6.

